# Understanding implementation of HEARTS for hypertension and diabetes in Guatemala: Qualitative and mixed-methods pilot results

**DOI:** 10.64898/2026.05.04.26352395

**Authors:** Taryn M. Valley, Chris Santizo-Malafronti, Irmgardt Alicia Wellmann, Luis Fernando Ayala, Nia Lucas, Mark D. Huffman, Anita Chary, Peter Rohloff, Rocío Donis, Ana Irina Argeli Alvarez Nufio, Manuel Ramírez-Zea, David Flood

**Affiliations:** School of Medicine and Public Health, University of Wisconsin-Madison, Madison, Wisconsin, USA; Department of Anthropology, University of Wisconsin-Madison, Madison, Wisconsin, USA; Department of Astronomy, University of Arizona, Tucson, Arizona, USA; INCAP Research Center for Prevention of Chronic Diseases, Institute of Nutrition of Central America and Panama, Guatemala City, Guatemala; University of Michigan Medical School, University of Michigan, Ann Arbor, Michigan, USA; Department of Medicine, School of Public Health, and Global Health Center, Washington University in St Louis, St Louis, Missouri, USA; The George Institute for Global Health, University of New South Wales, Sydney, Australia; Center for Indigenous Health Research, Wuqu’ Kawoq, Tecpán, Guatemala; Department of Emergency Medicine, Baylor College of Medicine, Houston, Texas, USA; Division of Global Health Equity, Brigham and Women’s Hospital, Boston, Massachusetts, USA; National Program for the Prevention of Chronic Non-Communicable Diseases and Cancer. Ministry of Health, Guatemala City, Guatemala; Department of Internal Medicine, University of Michigan, Ann Arbor, Michigan, USA

**Keywords:** hypertension, diabetes, implementation science, Guatemala, HEARTS, qualitative research, mixed methods, primary care, health systems, Latin America

## Abstract

**Objectives:** Most countries in Latin America have committed to adopting HEARTS, PAHO’s recommended approach for managing hypertension and diabetes in national primary care health systems. This study aimed to understand and refine a HEARTS pilot for scale-up in Guatemala.

**Methods:** Team members conducted semi-structured interviews with 30 patients and health workers participating in a HEARTS pilot across 10 primary care facilities in Guatemala’s Ministry of Health system. Researchers analyzed interviews using a combined inductive-deductive approach alongside the Tailored Implementation in Chronic Diseases framework and used convergent mixed methods to generate meta-inferences.

**Results:** Despite high feasibility and acceptability scores, health workers described tensions between HEARTS and competing responsibilities. Patients described rational navigation of an unreliable system, a more fitting explanation for low retention than noncompliance. Both patients and health workers understood HEARTS as another externally funded project with uncertain sustainability. Medication availability improved during the pilot, but district and facility-level incentive structures perpetuated supply unreliability. Greater absolute treatment gains for hypertension than for diabetes likely reflected health worker comfort and preexisting access to blood pressure monitoring supplies. Both patients and health workers identified education gaps. Integration of qualitative and quantitative findings suggested concrete scale-up refinements, including simplifying treatment protocols, strengthening diabetes components, focusing training on nurses, reforming central-level pharmaceutical supply policies, and securing high-level Ministry of Health commitments.

**Conclusions:** A HEARTS pilot trial in Guatemala met pre-specified quantitative outcomes, but qualitative and mixed-methods approaches revealed key barriers. These findings will assist in refining HEARTS in Guatemala for planned national scale-up.

**Trial registration:** ClinicalTrials.gov (NCT06080451)

## INTRODUCTION

Most people with hypertension or diabetes worldwide live in low- and middle-income countries (LMICs) (1). Yet only 30% of people with hypertension and 50% of people with diabetes in LMICs are treated; even fewer attain control (2, 3). The World Health Organization (WHO) and the Pan American Health Organization (PAHO) recommend the HEARTS primary care delivery model to close these implementation gaps. In Latin America, “HEARTS in the Americas” combines foundational modules of the WHO HEARTS Technical Package (4) and PAHO’s groundbreaking health system quality improvement tools (5). In Latin America, most countries have committed to adopting HEARTS, with substantial variation in health system implementation and institutionalization (6).

This study explores HEARTS implementation in Guatemala, a middle-income country and the most populous nation in Central America. Guatemala’s age- and population-adjusted prevalence of hypertension and diabetes exceeds regional peers, yet treatment levels are some of the region’s lowest (1, 7). Like many countries in the region and worldwide, Guatemala’s publicly funded, national Ministry of Health (MOH) system covers most (∼75%) of the population, providing an opportunity to produce generalizable knowledge on national health system HEARTS implementation (8).

We previously reported clinical and implementation outcomes from a single-arm HEARTS pilot trial in Guatemala (9). At the time of our pilot, the MOH of Guatemala had announced an agreement with PAHO to adopt HEARTS but had taken limited steps to advance implementation (10). The objective of this qualitative and mixed-methods study is to better understand the successes and challenges of our HEARTS pilot and to refine the model for planned national scale-up in Guatemala.

## MATERIALS AND METHODS

### Study design

We conducted a qualitative study consisting of semi-structured interviews with patients and health workers participating in a pilot feasibility and acceptability trial on HEARTS implementation in Guatemala (ClinicalTrials.gov NCT06080451). We followed the Consolidated Criteria for Reporting Qualitative Research (COREQ) guidelines in reporting results (11).

The trial protocol and quantitative results have been previously published (12, 13). The intervention aimed to improve the implementation of guideline-concordant primary care for patients with hypertension and/or diabetes and enrolled 964 patients with one or both of these conditions from October 2023 to May 2024 in 10 primary care facilities in the MOH system. The pilot’s primary outcomes were feasibility and acceptability, assessed through surveys with MOH employees and analysis of administrative data. These quantitative data suggested that HEARTS was feasible and acceptable. At the same time, we also observed suboptimal patient retention, low absolute treatment levels, and limited implementation fidelity for certain strategies.

### Implementation context

The pilot and these interviews took place in two Guatemalan MOH health districts: one serving a predominantly Indigenous population and the other a primarily non-Indigenous. Both districts were rural and characterized by high poverty rates (60-70%). MOH safety-net primary care is delivered through two community-level sites: health posts and health centers (8). Health posts operate in rural villages during weekday business hours, typically staffed by one or two auxiliary nurses, employed full-time by the MOH. Their training is comparable to nursing assistants in high-income countries. Before HEARTS implementation, health posts generally did not provide chronic disease services. Health centers, located in urban and semi-urban locations, operate approximately 8 hours daily. Their staff includes professional nurses (analogous to registered nurses), general physicians, and medical students. Patients with uncomplicated hypertension and diabetes are managed at this level using oral medications, while more complex patients are referred to regional or national hospitals.

The HEARTS pilot included five implementation strategies aligned with the WHO/PAHO HEARTS model (4). First, all primary health workers received training on standardized hypertension and diabetes clinical management protocols, including simplified medication initiation/titration algorithms. Second, district health authorities collaborated with external technical advisors to strengthen medicine procurement, inventory, and supply chains. Third, a team-based, task-sharing model expanded auxiliary and professional nurse roles, authorizing auxiliary nurses to initiate and titrate hypertension and diabetes medications. Fourth, an electronic monitoring tool was implemented. Fifth, health system administrators received feedback to guide quality improvement.

### Sampling and recruitment

For this study, we purposively sampled health workers and patients across health districts and sites because of their direct involvement and complementary perspectives on HEARTS implementation. We conducted 30 semi-structured interviews to achieve thematic saturation, informed by our prior qualitative studies in Guatemala and consistent with scholarly recommendations (14). Sampling aimed to achieve representation across gender, health district (Indigenous or non-Indigenous), and health worker role. Health workers were eligible if they worked at a participating facility during the HEARTS pilot with direct involvement in hypertension or diabetes care. Patient eligibility required care for hypertension and/or diabetes at a participating MOH facility during the pilot. Interviewers, field evaluators trained by the research team, identified potential participants who met sampling criteria and contacted them by phone or approached them in person to request an interview.

### Data collection

We developed two semi-structured interview guides. The patient interview guide focused on self-management practices, perceptions of health system quality, and community support. The health worker interview guides, tailored to specific role, focused on clinical responsibilities, experiences with each HEARTS strategy, implementation determinants per the Tailored Implementation in Chronic Diseases (TICD) checklist (15), and suggestions for HEARTS refinement. District managers were asked about system-level TICD items, including budget, human resources, and organizational leadership.

The research team scheduled interview timing directly with participants. Patient interviews were conducted in patients’ homes, and health worker interviews in a private location at the health facility. The research team selected and trained interviewers based on their professional experience and fluency in both Spanish and Kaqchikel, the Mayan language in the Indigenous health district; one interviewer was a man and one a woman. TranscribeMe, a professional transcription service, transcribed Spanish-language interviews. A professional linguist transcribed Kaqchikel interviews and simultaneously generated Spanish translations. Audio recordings allowed researchers to verify transcripts.

### Qualitative data analysis

Two master’s-level qualitative researchers (TMV and CS-M) coded the Spanish transcripts. Both were fluent in Spanish and English, and TMV had advanced proficiency in Kaqchikel. We used a combined inductive-deductive approach, drawing from implementation scientists who emphasize fit and stakeholder engagement when determining qualitative methodology (16). We prioritized immediately actionable content and HEARTS-related themes. For health worker interviews, we used the TICD checklist (15) to generate initial codes, added emergent codes, and focused on themes spanning professional roles. For patient interviews, we primarily used emergent codes but prioritized relevance to HEARTS.

Transcripts were coded in Dedoose (Version 10.0.59, 2026). When the two researchers had consensus-coded three patient and four health worker transcripts, they felt confident applying the codebook independently. During the independent phase of coding, when questions or proposals for new codes arose, the coders met to make joint decisions. Most codes remained stable from initial codebook to final analysis. TMV read all coding reports to generate themes. The team finalized themes through discussion and revisiting coding reports.

During the preparation of this work, the authors used Anthropic Claude Opus 4.6 (claude.ai) as an iterative writing and copyediting tool. AI was not used in the qualitative analysis, and no figures or wholesale text were AI-generated.

### Mixed-methods integration of quantitative and qualitative data

We integrated quantitative findings from the pilot trial (13) and qualitative findings from semi-structured interviews using convergent mixed methods (17). Two team members (TMV and DF) organized qualitative and quantitative findings by theme, drawing meta-inferences about HEARTS implementation.

## RESULTS

### Demographic characteristics of participants

The sample consisted of 30 participants, 20 health workers and 10 patients. Ten auxiliary nurses, four professional nurses, four physicians, and two district managers comprised the health worker sample, representing 10 facilities across the two study districts. Five patients participated from each health district, six of ten were women, with mean patient age 61 years.

Six patients were interviewed in Spanish, and four in Kaqchikel. Eight patients had received hypertension and/or diabetes care before enrolling in HEARTS.

**Table 1** describes patient and provider perspectives on HEARTS pilot implementation, notated by quotation number in the text.

**Table 1:**
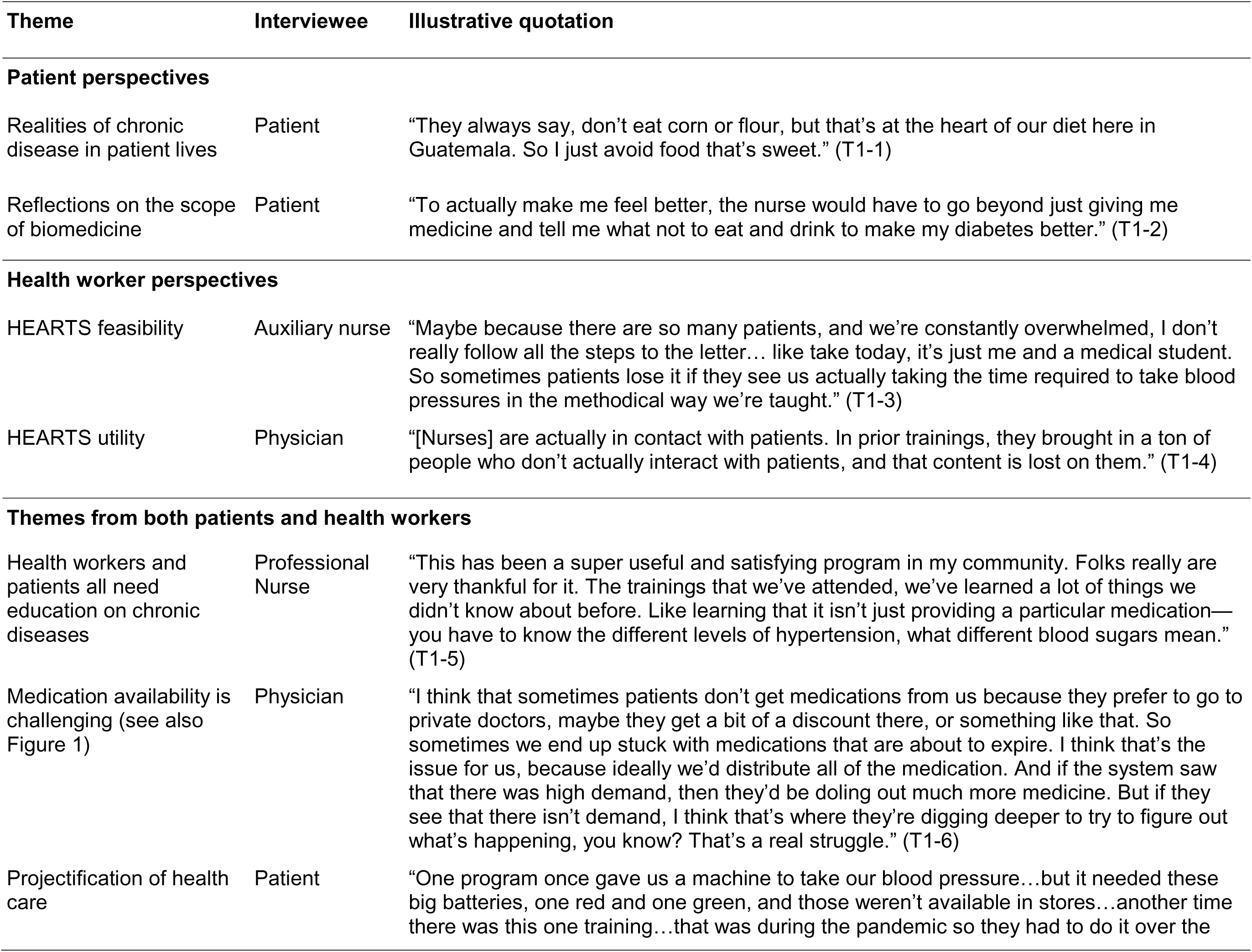
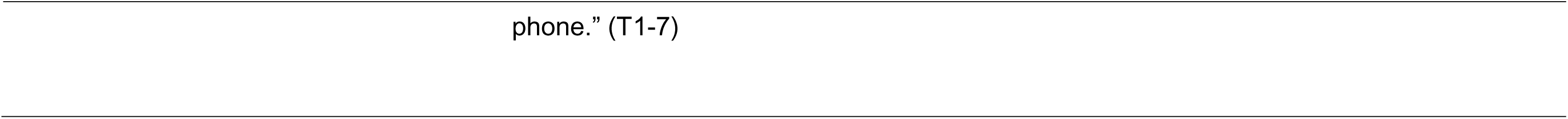
Patient and provider perspectives on HEARTS pilot implementation.

| Theme | Interviewee | Illustrative quotation |
| --- | --- | --- |
| **Patient perspectives** |  |  |
| Realities of chronic disease in patient lives | Patient | “They always say, don’t eat corn or flour, but that’s at the heart of our diet here in Guatemala. So I just avoid food that’s sweet.” (T1-1) |
| Reflections on the scope of biomedicine | Patient | “To actually make me feel better, the nurse would have to go beyond just giving me medicine and tell me what not to eat and drink to make my diabetes better.” (T1-2) |
| **Health worker perspectives** |  |  |
| HEARTS feasibility | Auxiliary nurse | “Maybe because there are so many patients, and we’re constantly overwhelmed, I don’t really follow all the steps to the letter... like take today, it’s just me and a medical student. So sometimes patients lose it if they see us actually taking the time required to take blood pressures in the methodical way we’re taught.” (T1-3) |
| HEARTS utility | Physician | “[Nurses] are actually in contact with patients. In prior trainings, they brought in a ton of people who don’t actually interact with patients, and that content is lost on them.” (T1-4) |
| **Themes from both patients and health workers** |  |  |
| Health workers and patients all need education on chronic diseases | Professional Nurse | “This has been a super useful and satisfying program in my community. Folks really are very thankful for it. The trainings that we’ve attended, we’ve learned a lot of things we didn’t know about before. Like learning that it isn’t just providing a particular medication—you have to know the different levels of hypertension, what different blood sugars mean.” (T1-5) |
| Medication availability is challenging (see also Figure 1) | Physician | “I think that sometimes patients don’t get medications from us because they prefer to go to private doctors, maybe they get a bit of a discount there, or something like that. So sometimes we end up stuck with medications that are about to expire. I think that’s the issue for us, because ideally we’d distribute all of the medication. And if the system saw that there was high demand, then they’d be doling out much more medicine. But if they see that there isn’t demand, I think that’s where they’re digging deeper to try to figure out what’s happening, you know? That’s a real struggle.” (T1-6) |
| Projectification of health care | Patient | “One program once gave us a machine to take our blood pressure...but it needed these big batteries, one red and one green, and those weren’t available in stores...another time there was this one training...that was during the pandemic so they had to do it over the |

### Themes elicited from patients

#### Chronic disease realities

Patients described how Guatemalan nutrition education about diabetes often mandates avoiding sugary food; patients may then cut out food that tastes sweet, without limiting carbohydrates overall. Health workers and patients took for granted diets reliant on culturally important, simple, savory carbs, like atol (a nutrient-rich corn-based drink), corn tortillas, and white bread (T1-1).

Beyond diet, patients attempted to make sense of their illnesses, sometimes in ways at odds with HEARTS. Some patients worried that high blood pressure caused diabetes or feared that diabetes stemmed from medications. One patient explained that, though he took his medication, he was unsure how to control his diabetes:

> “Honestly, they haven’t even told me how I’m going to actually get ahead of this diabetes. Sure, I need to take my medicines, I’m taking them, but they haven’t told me how I’m going to, shall we say, get rid of the diabetes itself.”

Like other interviewees, this patient suggested that medicine could “get rid of” diabetes, echoing curative language one might use for a viral infection or other time-limited disease. These and other comments exemplified underlying misalignments between the biomedical view of hypertension and diabetes pathophysiology, as espoused by HEARTS, and patients’ conceptual models. Such divergent understandings shaped patients’ expectations about disease trajectory and appropriate treatment.

#### Biomedicine’s scope in patients’ lives

While some patients received diet or exercise education, others noted that medicines seemed the main, or only, health system intervention (T1-2). Several described wariness about taking pharmaceuticals, what many called “chemical” medicines (contrasted with “natural” or traditional medicines). One patient explained, “Sometimes I find myself thinking, how is it possible that taking all these pills won’t hurt me?” Nonetheless, she reported taking her medicine regularly, wishing that “these medicines were easier to get at the health center.”

Some patients commented on low-quality medical care. One patient’s daughter, who worked at the MOH herself, explained that:

> “Sometimes at the health center, who knows why, you get there, and the staff is in a super bad mood, and they aren’t willing to actually listen to every patient. So my mom decided not to go back at all.”

#### HEARTS within an unreliable health system

Patients reported cobbling together advice from physicians, family members, and neighbors to determine where to seek chronic disease care. One patient, exemplifying creativity consistent across patients, sometimes purchased medicine from a private physician, other times purchased medication directly from a pharmacy, but always prioritized “finding medication they’re giving away for free,” as through HEARTS. Another patient, prior to HEARTS, sought care in different places for diabetes versus hypertension: the original health center that diagnosed her high blood pressure “didn’t have the tools to check blood sugar,” so she was instructed “to go find a private doctor and check my sugars there.” Another patient continued to go to the MOH center for hypertension medications, but saw a private doctor for diabetes.

### Themes elicited from health workers

Health workers described significant heterogeneity of chronic disease care at health centers and posts. One health post had one registered diabetes patient; another health post reported 15 registered patients but estimated 80 patients including unregistered. The number of reported employees at facilities ranged from three to over 100. Most health facilities saw diabetes and hypertension patients monthly, though one facility saw patients weekly and another every three months. Three of 10 health posts offered diabetes support groups.

#### HEARTS feasibility and utility

Health workers explained that, in practice, implementing HEARTS as they were taught conflicted with clinical realities. While some health workers said HEARTS strategies were generally feasible, others described that regular understaffing, alongside patients’ frustrations about long waits, meant rarely taking blood pressures using exact HEARTS procedures (T1-3). A professional nurse explained:

> “There’s a ton of pressure from the Ministry: Getting what they deem as adequate vaccine coverage, getting antiparasitics and supplementation to kids under 5, vaccinating women…those are the programs the Ministry actually prioritizes… It’s really hard to do additional tasks, above and beyond those, even though the Ministry theoretically expects us to do it all.”

When employees cannot manage even the tasks they have been told are necessary and prioritized, HEARTS seems both lower priority and less feasible.

Health workers emphasized that HEARTS utility depended on capacity to work within the sometimes counterintuitive processes determining day-to-day activities at MOH facilities. Some felt that incorporating electronic and physical registries captured information that “otherwise we wouldn’t even ask about; that stuff gets away from us.” A different physician said chronic diseases had historically been “abandoned by the Ministry,” so HEARTS pilot trainings helped staff refocus on hypertension and diabetes. One doctor requested training target nurses (T1-4). Several commented that staff working in medication supply and ordering rarely communicated directly with clinical staff, contributing to medication access issues.

### Themes elicited from both patients and health workers

#### Education

Both patients and health workers thought education, critical to HEARTS implementation, could be strengthened. Health workers needed training from the MOH to effectively care for diabetes and hypertension patients who, in turn, needed clearer education about their conditions. Many health workers said trainings directly improved the care they provided and asked for additional training (T1-5).

#### Medication availability

Patients and health workers highlighted medication access as the most salient of many resource limitations within the MOH. Patients at almost every health post cited medication shortages; one patient explained how “they say, we don’t have that medicine right now, all we have is this other one,” which that patient did not receive.

Patients rarely blamed local MOH staff for medication shortages. The same patient explained succinctly: “Health workers have the best intentions, but the supply chain doesn’t work.” Another patient saw medication provision as the cornerstone of her expectations of the MOH:

> “What we actually want is for them to give us all of our medications. Sometimes there’s no medicine, and they don’t give us any. What we need is for there to be medicine available and for them to provide good care.”

Some health workers attributed low medication demand to patients going outside the MOH system (T1-6). Patients explained perceived higher-quality care in the private sector and MOH stockouts. Health workers described an MOH policy mandating that frontline MOH staff, mostly nurses, must reimburse the MOH out of their personal salary for any unused or expired medication. In contrast, if all medication is distributed, health workers do not face a penalty. As a consequence, MOH employees, despite the best of intentions, are incentivized to prefer to turn patients away without medication rather than have medication remaining at the end of the supply period. See **Figure 1** for a visual depiction of this feedback loop.

**Figure 1:**
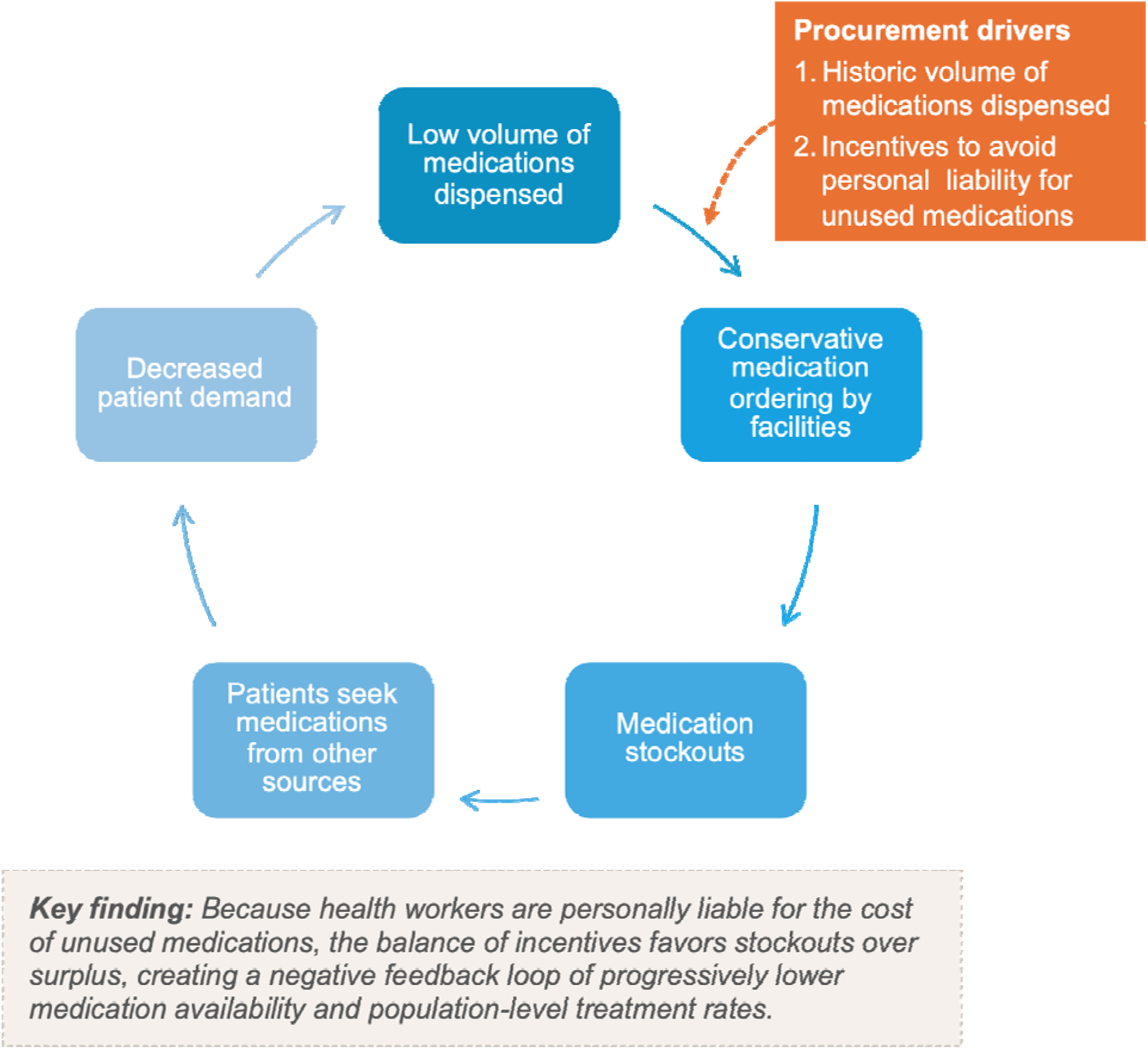
**Vicious cycle of demand-based medication procurement**

#### Projectification of health

Both patients and health workers expressed understanding that getting medical care in current-day Guatemala required patients to navigate outside-funded, often short-term, inherently unreliable projects. This demanded patient creativity, and interviewees understood care to be projectified (T1-7).

Amidst chronic project cycles, MOH employees hoped the HEARTS program would remain. One auxiliary nurse asked interviewees, “Well, is the program going to end, or is it going to keep going?” The interviewer responded, “There’ll be follow-up, like in a different type of project, through training. But in its current format, the HEARTS pilot is wrapping up at this point.” The nurse summarized:

> “Yeah, it’d be good if there were an opportunity for us to be trained and all of that… there are new and different things every day. So in order for us to stay up-to-date, that would be a benefit, and to be able to improve our work with patients.”

### Findings from mixed-methods integration of quantitative and qualitative data

**Table 2** shows the joint display integrating quantitative, qualitative, and mixed methods meta-inferences. Despite high feasibility and acceptability scores, health workers described tensions between HEARTS protocols and competing responsibilities. Low patient retention (36% at 3 months) reflected rational navigation of an unreliable system rather than noncompliance. Both patients and health workers understood HEARTS as another externally funded, potentially temporary initiative, creating uncertainty despite high sustainability scores (4.7 to 5.7 on a 7-point scale). Medication availability improved (60% to 81% of facilities), but a critical barrier emerged: health workers were personally charged for expired medications, incentivizing under-ordering and perpetuating stockouts (**Figure 1**). Hypertension showed greater treatment gains than diabetes (22.3 vs. 3.5 patients per month increase), reflecting greater health worker comfort and available blood pressure management tools. Finally, both patients and health workers identified education as a critical area to strengthen.

**Table 2:** Mixed-methods joint display showing integration of quantitative and qualitative findings.

| Domain | Quantitative findings | Qualitative findings | Mixed methods integration | Implications for refinement |
| --- | --- | --- | --- | --- |
| *Primary outcomes* |  |  |  |  |
| Feasibility and acceptability | Feasibility and acceptability surveys among health workers had median scores of 5.0 (IQR: 5.0 to 5.0) and 5.0 (IQR: 4.8 to 5.0), respectively (higher is better), exceeding benchmark of 3.5 | While health workers perceived HEARTS as generally feasible and acceptable and important, it was impossible to follow certain steps due to the time burden<br>Competing MOH priorities | Despite high survey scores, health workers described tensions adhering to HEARTS protocols given demands of other programs | Further simplify HEARTS strategies<br>More explicitly consider competing priorities of health workers |
| Patient enrollment | Enrolled 964 patients, exceeding benchmark of 25 patients per district | Strong interest among patients and health workers to improve HTN and DM care | Quantitative and qualitative data converged to show demand for HEARTS in the MOH system | Ground-level stakeholders (patients and health workers) support HEARTS |
| Patient retention | Only 36% of patients attended a follow-up visit within 3 months, below the benchmark of 75% | Patients rationally navigated an unreliable health system<br>Medications a clear driver of low retention in care | Low retention reflected system unreliability, not patient non-compliance or disinterest | Must improve systemic issues of medication supply, consistent staffing, and care quality |
| *Secondary outcomes* |  |  |  |  |
| Treatment rates | HTN treatment rates increased by 22.3 patients per month (95% CI: 16.2 to 28.4); DM treatment rates increased by 3.5 patients per month (95% CI: -1.6 to 8.7) | Greater health worker comfort managing HTN than DM<br>DM supplies such as glucometers unreliable, contributing to DM challenges | Health worker comfort and resource availability likely explain HTN vs DM gap | Strengthen diabetes training and ensure reliable diabetes monitoring equipment and supplies as HEARTS scales up in Guatemala |
| Disease control | Among patients with data, 50.6% achieved BP control and 41.0% achieved glycemic control; baseline therapeutic intensity was low (HTN: 0.8, DM: 1.2) | Health workers did not raise the issue of treatment intensity<br>Patient confusion about the concept of control vs. cure | There were specific key gaps in education about HTN and DM among both health workers and patients | Strengthen education on:<br>Health worker understanding of treatment intensity<br>Patient understanding of goal in managing chronic conditions |
| Sustainability | Sustainability scores among health workers ranged from 4.7 to 5.7 on 7-point scale (higher is better) | Health workers and patients raised sustainability questions given history of temporary external programs in the MOH | Positive perception of theoretical sustainability balanced by a degree of stakeholder skepticism | HEARTS institutionalization requires high-level MOH support/commitment beyond previous ground-level initiatives |
| *Implementation strategies* |  |  |  |  |
| Health worker training | 20 workers per district attended all training sessions | Training highly valued<br>Requested ongoing sessions<br>Suggested targeting frontline clinical staff, especially nurses<br>Sought more diabetes training | Health workers wanted ongoing reinforcement and better targeting to nurses, especially on diabetes topics | HEARTS training is excellent, but scale-up should prioritize nurses and diabetes to a greater degree than in the pilot |
| Team-based care | Nurses prescribed 57% of HTN and | Clinical task-sharing as | Task-sharing succeeded clinically | Unrealistic to expect monthly in- |
| | DM medicines; only 10% of facilities conducted $\geq 1$ care coordination meeting per month | recommended in HEARTS was perceived as important<br>Time, cultural, and organizational constraints limited multidisciplinary meetings | with nurses writing majority of prescriptions, but multidisciplinary meetings not held due to structural constraints | person meetings among nurses and physicians, so asynchronous coordination options are needed |
| Medication and diagnostic access | Medication availability improved from 60% to 81% of facilities; core antihypertensives were available at 82% of facilities and core diabetes medications at 80% | Medications were most critical barrier to HEARTS success<br>Health worker salary penalty for expired medications | MOH policy creates rational under-ordering and stockouts | Centralize drug purchasing<br>Remove staff's personal financial penalty for expired medications |
| Electronic monitoring | Only 7.6% of patient visits were captured in the electronic monitoring system; usability score was 67.7 on 100-point scale (higher is better) | Health workers required $\geq 20$ minutes charting per patient<br>Many competing documentation demands | Time burden incompatible with resource context | Simplify electronic system and streamline processes of data entry |

Abbreviations: BP: blood pressure; DM: diabetes mellitus; HTN: hypertension; MOH: Ministry of Health; Qual: qualitative; Quant: quantitative.

## DISCUSSION

This study aimed to understand the mixed success of a HEARTS implementation pilot trial in the national public primary care system in Guatemala. Pilot study qualitative and mixed methods approaches complemented quantitative trial results (13), yielding refinements for scale-up. First, we found that HEARTS strategies and protocols in Guatemala should be simplified, given health worker competing priorities. Second, improving patient retention requires structural health system changes around drug supply, staffing, and improving care quality and capacity—rather than focusing on changing the behavior of individual patients or individual health workers. Third, diabetes components need greater emphasis compared to hypertension, especially in training and supply provision. Fourth, training should prioritize nurses to a greater degree, as nursing involvement is indispensable to expanding treatment in HEARTS. Fifth, MOH policy changes are needed to reform pharmaceutical supply chains and eliminate punitive incentive structures that perpetuate stockouts.

This pilot study has implications for improving hypertension and diabetes care in the nearly 40 countries that have adopted HEARTS, including dozens of Latin American countries with similar health systems. Regarding drug access, PAHO recognizes medication availability as a critical HEARTS barrier (18) and has recommended revising national essential medicine lists, prioritizing fixed-dose combinations, and limiting the number of unique therapeutic agents and dose sizes in formularies (19). This study illustrates the complex, multilevel nature of pharmaceutical barriers. Beyond national regulations or policy, incentive structures operating at primary care facility levels shape medication availability in ways that are observable even in the daily work of individual health workers. We term this observation a “vicious cycle of demand-based medication procurement” (see **Figure 1**). The result is very low population-level treatment rates; approximately 1% of people with hypertension or diabetes receive treatment from the MOH at a given time (20).

Roles of non-physician health workers as part of the team-based care provides another area for generalization. In this pilot, nurses prescribed nearly 60% of hypertension and diabetes medications, despite virtually no formal coordination between physicians and nurses due to time, cultural, and organizational constraints. Guatemala’s health system, as other countries’, historically has been physician-centered (21). Our results indicate the importance of formal regulations and policies facilitating nurse medication prescribing and titrating within MOH-standardized algorithms—without requiring physician supervision or approval. This HEARTS approach has shown promise in El Salvador and Chile (21).

This study reveals tension between simplicity and completeness in HEARTS implementation. HEARTS has innovatively bundled straightforward, synergistic hypertension interventions: simplified protocols, minimal drug formularies, and task shifting. PAHO’s “HEARTS 2.0” aims to add more interventions covering multiple cardiometabolic risk factors, including diabetes, chronic kidney disease, and dyslipidemia (9). We have previously argued integrating management of these risks, especially diabetes (22). A holistic approach aligns with how this study’s patients conceptualized their health; they cared about the total effect of illnesses on their lives, not a specific disease or a single cardiometabolic risk factor. However, countries adopting HEARTS must weigh the trade-offs between program comprehensiveness and health system capacity. Latin America’s readiness for HEARTS 2.0 varies widely by country (23). In this study, health workers repeatedly emphasized the need for maximally simple and efficient HEARTS strategies, given extraordinary competing demands on their time.

This qualitative and mixed methods study focused on ground-level stakeholders (patients, health workers, and district managers), not higher-level MOH and government officials: we address this gap in forthcoming HEARTS research. In 2024, after the pilot concluded, the newly elected presidential administration brought increased political commitment to strengthening MOH primary care in three actions: First, the administration established an agreement with the United Nations Office for Project Services to centralize medication and supply procurement, valued at up to $900 million over four years (approximately 35% of MOH procurement) (24).

This approach, though not through the PAHO Strategic Fund, aligns with HEARTS recommendations for streamlined national procurement. But this new centralized procurement approach has faced opposition from entrenched private pharmaceutical sector interests (25). Second, the Presidential administration announced plans to expand the Medicine Access Program, a government-operated drug revolving fund providing low-cost medications through municipal pharmacies (26). While not eliminating patient out-of-pocket costs entirely, such mechanisms have shown promise in other countries’ large-scale HEARTS programs (27). In Guatemala, however, municipal-led drug revolving funds are at risk of becoming politicized, threatening their sustainability when sponsoring political parties lose power (28). Third, in one of the Guatemalan states where the pilot was conducted, the Presidential administration signed an agreement to integrate traditional Indigenous medicine into the MOH (29). Our group will seek to investigate the effects of these policies in future research.

This project’s findings point to a broader challenge in global implementation science: the tendency of frameworks to locate barriers at the level of individual actors with limited agency, rather than at the level of systems operating under immense structural constraints. Recent scholarship has called for the field to more fully consider power’s role in shaping program implementation and research agendas (30). Frameworks developed in high-income contexts, like the Expert Recommendations for Implementing Change (ERIC) compilation and the Consolidated Framework for Implementation Research (CFIR), have been critiqued for incompletely addressing system- and policy-level factors in LMICs (31, 32). In this study, we used the TICD checklist to guide qualitative analysis because it includes a system-level domain addressing health system budget constraints, corruption, influential people, political stability, and other barriers (15). We found that both patients and health workers understood HEARTS as an externally funded project with uncertain sustainability, creating what medical anthropologists term an “enclave” of projectified care (33). Patients engaged in “therapeutic clientship” (34)—strategically accessing care by navigating between temporary, externally funded projects— instead of consistency through the MOH. Simultaneously, health workers could not adhere to HEARTS strategies because of impossible workloads, not lack of skill or interest. Through this lens, retention in HEARTS was not a patient-level barrier requiring behavioral interventions but rather a rational manifestation of patients seeking care under structural constraints. Global implementation research must resist laundering system-level failures into deficits of patients and frontline workers.

This study has limitations. First, this was a relatively small study among patients and health workers sampled purposefully rather than probabilistically. This study design was appropriate for our objective of understanding pilot results. Second, the underlying pilot trial was not randomized, lasted only six months, and did not collect patient-level disease control data. We are currently planning a fully powered randomized trial with longer follow-up, which will strengthen mixed-methods integration by allowing us to triangulate qualitative barriers with clinically relevant causal endpoints. Third, we focused on ground-level stakeholders and did not interview high-level Ministry of Health administrators, politicians, PAHO officials, or pharmaceutical representatives, while HEARTS is fundamentally a health system intervention relying on political, policy, and funding decisions at elite levels. Fourth, the pilot trial ended in May 2024, before significant political developments in Guatemala, and enhancements in PAHO’s “HEARTS in the Americas” initiative (9, 35). This paper’s findings thus may not fully capture present MOH HEARTS implementation dynamics. Finally, health workers may have felt pressure to report positive experiences with HEARTS, and patients may have moderated criticisms out of concern about jeopardizing their care or HEARTS continuation. We believe the risk of social desirability bias is relatively small: the research team was from INCAP, a highly respected institution among Guatemalans. Our experience is that patients generally recognize that the MOH (not INCAP) is responsible for delivering health care.

## CONCLUSIONS

This HEARTS pilot in Guatemala met quantitative feasibility and acceptability outcomes, but qualitative and mixed-methods approaches revealed important implementation barriers. This paper’s qualitative and mixed methods findings have informed refinements to the HEARTS model as Guatemala’s Ministry of Health proceeds with national scale-up, including simplifying treatment protocols, strengthening diabetes components, reforming pharmaceutical supply policies, and securing high-level political commitments.

## Supporting information

COREQ checklist

## SUPPLEMENTARY INFORMATION AND DECLARATIONS

### Supplementary material

Consolidated Criteria for Reporting Qualitative Research (COREQ) reporting guidelines

## Acknowledgements

We wish to acknowledge the efforts of Ministry of Health staff, clinicians, and patients who are involved in the HEARTS program in Guatemala.

## Authors’ contributions

DF conceived the idea for this study and obtained funding. IAW and LFA led data collection, under the supervision of DF and MR-Z. TMV and CS-M coded the interviews with assistance from DF. TMV and DF wrote the first draft of the manuscript with substantial revisions from the rest of the author team. All authors approved this submission.

## Availability of data and materials

Statistical code, data dictionaries, and training materials from the pilot trial are publicly available on the Harvard Dataverse (https://doi.org/10.7910/DVN/0X8QKE). Certain quantitative datasets will be made available upon reasonable request to the authors; however, some datasets include Ministry of Health administrative data that cannot be shared. Due to participant privacy concerns and risk of re-identification, we do not plan to share qualitative data such as audio recordings or transcripts.

## Declaration of AI Use

During the preparation of this work, the authors used Anthropic Claude Opus 4.6 (claude.ai) as an iterative writing and copyediting tool. AI was not used in the qualitative analysis, and no figures or wholesale text were AI-generated. After using this tool, all authors reviewed and edited the content as needed and take full responsibility for the content of the publication.

## Ethics approval and consent to participate

This research was approved by the ethics committees of the Ministry of Health of Guatemala (07–2023), the Institute of Nutrition of Central America and Panama (CIE-REV 124/2023), and the University of Michigan (HUM00234613). Verbal informed consent was obtained from patients and health workers for their participation in semi-structured interviews.

## Consent for publication

Not applicable.

## REFERENCES

1. Zhou B, Carrillo-Larco RM, Danaei G, Riley LM, Paciorek CJ, Stevens GA, et al. Worldwide trends in hypertension prevalence and progress in treatment and control from 1990 to 2019: a pooled analysis of 1201 population-representative studies with 104 million participants. The Lancet. 2021.

2. Geldsetzer P, Manne-Goehler J, Marcus ME, Ebert C, Zhumadilov Z, Wesseh CS, et al. The state of hypertension care in 44 low-income and middle-income countries: a cross-sectional study of nationally representative individual-level data from 1.1 million adults. Lancet. 2019 Aug 24;394(10199):652–62. PubMed PMID: 31327566. Epub 2019/07/23.

3. Flood D, Seiglie JA, Dunn M, Tschida S, Theilmann M, Marcus ME, et al. The state of diabetes treatment coverage in 55 low-income and middle-income countries: a cross-sectional study of nationally representative, individual-level data in 680 102 adults. Lancet Healthy Longev. 2021 Jun;2(6):e340–e51. PubMed PMID: 35211689. PMCID: PMC8865379. Epub 2022/02/26.

4. WHO. Hearts: Technical package for cardiovascular disease management in primary health care. Geneva: World Health Organization; 2016.

5. PAHO. HEARTS en las Américas: Marco de evaluación para la mejora continua de la calidad en los centros de atención primaria de salud. Washington, D.C.: Pan American Health Organization; 2024.

6. Ordunez P, Campbell NRC, DiPette DJ, Jaffe MG, Rosende A, Martinez R, et al. HEARTS in the Americas: Targeting Health System Change to Improve Population Hypertension Control. Curr Hypertens Rep. 2024 Apr;26(4):141–56. PubMed PMID: 38041725. PMCID: PMC10904446. Epub 20231202.

7. N. C. D. Risk Factor Collaboration. Worldwide trends in diabetes prevalence and treatment from 1990 to 2022: a pooled analysis of 1108 population-representative studies with 141 million participants. Lancet. 2024 Nov 23;404(10467):2077–93. PubMed PMID: 39549716. PMCID: PMC7616842. Epub 20241113.

8. Avila C, Bright R, Gutierrez J, Hoadley K, Manuel C, Romero N, et al. Guatemala Health System Assessment, August 2015. Bethesda, MD: Health Finance & Governance Project, Abt Associates Inc.; 2015.

9. Rosende A, Romero C, DiPette DJ, Brettler J, Van der Stuyft P, Satheesh G, et al. Candidate Interventions for Integrating Hypertension and Cardiovascular-Kidney-Metabolic Care in Primary Health Settings: HEARTS 2.0 Phase 1. Global heart. 2025;20(1).

10. Pan American Health Organization. Implementarán iniciativa Hearts para la prevención y el control de las enfermedades cardiovasculares (ECV) en Guatemala 2022 [Available from: https://www.paho.org/es/noticias/11-11-2022-implementaran-iniciativa-hearts-para-prevencion-control-enfermedades.

11. Tong A, Sainsbury P, Craig J. Consolidated criteria for reporting qualitative research (COREQ): a 32-item checklist for interviews and focus groups. International journal for quality in health care : journal of the International Society for Quality in Health Care / ISQua. 2007 Dec;19(6):349–57. PubMed PMID: 17872937. Epub 20070914.

12. Wellmann IA, Ayala LF, Rodriguez JJ, Guetterman TC, Irazola V, Palacios E, et al. Implementing integrated hypertension and diabetes management using the World Health Organization’s HEARTS model: protocol for a pilot study in the Guatemalan national primary care system. Implement Sci Commun. 2024 Jan 9;5(1):7. PubMed PMID: 38195600. PMCID: PMC10775666. Epub 20240109.

13. Wellmann IA, Ayala LF, Valley TM, Irazola V, Huffman MD, Heisler M, et al. Evaluating the World Health Organization’s Hearts Model for Hypertension and Diabetes Management: A Pilot Implementation Study in Guatemala. Global heart. 2025;20(1):9. PubMed PMID: 39896314. PMCID: PMC11784498. Epub 20250131.

14. Francis JJ, Johnston M, Robertson C, Glidewell L, Entwistle V, Eccles MP, et al. What is an adequate sample size? Operationalising data saturation for theory-based interview studies. Psychol Health. 2010 Dec;25(10):1229–45. PubMed PMID: 20204937. Epub 2010/03/06.

15. Flottorp SA, Oxman AD, Krause J, Musila NR, Wensing M, Godycki-Cwirko M, et al. A checklist for identifying determinants of practice: a systematic review and synthesis of frameworks and taxonomies of factors that prevent or enable improvements in healthcare professional practice. Implementation science : IS. 2013 Mar 23;8:35. PubMed PMID: 23522377. PMCID: PMC3617095. Epub 2013/03/26.

16. Ramanadhan S, Revette AC, Lee RM, Aveling EL. Pragmatic approaches to analyzing qualitative data for implementation science: an introduction. Implement Sci Commun. 2021 Jun 29;2(1):70. PubMed PMID: 34187595. PMCID: PMC8243847. Epub 20210629.

17. Guetterman TC, Fetters MD, Creswell JW. Integrating Quantitative and Qualitative Results in Health Science Mixed Methods Research Through Joint Displays. Ann Fam Med. 2015 Nov;13(6):554–61. PubMed PMID: 26553895. PMCID: PMC4639381. Epub 2015/11/11.

18. DiPette DJ, Skeete J, Ridley E, Campbell NRC, Lopez-Jaramillo P, Kishore SP, et al. Fixed-dose combination pharmacologic therapy to improve hypertension control worldwide: Clinical perspective and policy implications. J Clin Hypertens (Greenwich). 2019 Jan;21(1):4–15. PubMed PMID: 30480368. Epub 2018/11/28.

19. Souza KM, Giron N, Vallini J, Hallar K, Ordunez P, Rosende A, et al. Barriers to access to antihypertensive medicines: insights from the HEARTS initiative in latin American and Caribbean region. Journal of pharmaceutical policy and practice. 2024;17(1):2379045. PubMed PMID: 39109358. PMCID: PMC11302458. Epub 20240805.

20. Wellmann IA, Rodriguez JJ, Batzin B, Hegel G, Ayala LF, Ozano K, et al. Reach and effectiveness of a HEARTS hypertension pilot project in Guatemala. Revista panamericana de salud publica = Pan American journal of public health. 2024;48:e100. PubMed PMID: 39431198. PMCID: PMC11488152. Epub 20241018.

21. Irazola V, Prado C, Rosende A, Flood D, Tsuyuki R, Neira Ojeda C, et al. Expanding team-based care for hypertension and cardiovascular risk management with HEARTS in the Americas. Revista Panamericana de Salud Pública. 2025;49.

22. Flood D, Edwards EW, Giovannini D, Ridley E, Rosende A, Herman WH, et al. Integrating hypertension and diabetes management in primary health care settings: HEARTS as a tool. Revista panamericana de salud publica = Pan American journal of public health. 2022;46:e150. PubMed PMID: 36071915. PMCID: PMC9440730. Epub 2022/09/09.

23. Ordunez P, Rosende A, Brettler J, Londono E, Van der Stuyft P, Martinez-Piedra R, et al. Readiness to deliver integrated cardiovascular, kidney and metabolic care in primary healthcare: phase II of HEARTS 2.0 in 26 countries in the Americas. BMJ Glob Health. 2026 Jan 14;11(1). PubMed PMID: 41534947. Epub 20260114.

24. MSPAS, UNOPS. Ficha Técnica del Proyecto: “Mejora del acceso a la salud, por medio de alternativas para el aprovisionamiento de medicamentos, tecnología médica y otros insumos y para la red pública de salud de Guatemala”: Ministerio de Salud Pública y Asistencia Social (MSPAS and Oficina de las Naciones Unidas de Servicios para Proyectos (UNOPS); 2024 [Available from: https://storage.googleapis.com/unops-proyectos-guatemala/images/Ficha-Te%CC%81cnica-del-Proyecto.pdf.

25. Marroquín CP, Vargas E, Domínguez A. Embargan salario del ministro Joaquín Barnoya y suspenden pagos a UNOPS por orden judicial: Prensa Libre; 2026 [Available from: https://www.prensalibre.com/guatemala/justicia/embargan-salario-del-ministro-joaquin-barnoya-y-suspenden-pagos-a-unops-por-orden-judicial-breaking/.

26. Alonzo C. Meta presidencial, proyectan 12 nuevas grandes farmacias Proam: Agencia Guatemalteca de Noticias; 2025 [Available from: https://agn.gt/meta-presidencial-proyectan-12-nuevas-grandes-farmacias-proam/.

27. Shedul GJ, Ugwuneji EN, Jamro EL, Banigbe B, Ponzing P, Okpe I, et al. Design and implementation of a drug revolving fund for hypertension treatment in primary care setting of the Federal Capital Territory, Nigeria. BMJ Glob Health. 2025 Nov 19;10(11). PubMed PMID: 41265927. PMCID: PMC12636893. Epub 20251119.

28. Wellmann Castellanos IA, Hegel G, Rodríguez DJ, Ayala LF, Ramirez-Zea M, Flood D, editors. Revolving fund pharmacies: An implementation strategy for the World Health Organization’s “HEARTS” model of cardiovascular disease management in primary care. 16th Annual Conference on the Science of Dissemination and Implementation in Health; 2023; Arlington, VA: Academy Health.

29. Ministerio de Desarrollo Social de Guatemala. Ministro firma Pacto Nacional de Salud: MIDES; 2024 [Available from: https://www.mides.gob.gt/ministro-firma-pacto-nacional-de-salud/.

30. Fischer HT, Hanefeld J, Koduah A. Implementation science and power: equity-oriented implementation science needs a power lens. The Lancet Global health. 2025 Nov 11. PubMed PMID: 41237805. Epub 20251111.

31. Means AR, Kemp CG, Gwayi-Chore MC, Gimbel S, Soi C, Sherr K, et al. Evaluating and optimizing the consolidated framework for implementation research (CFIR) for use in low- and middle-income countries: a systematic review. Implementation science : IS. 2020 Mar 12;15(1):17. PubMed PMID: 32164692. PMCID: PMC7069199. Epub 2020/03/14.

32. Lovero KL, Kemp CG, Wagenaar BH, Giusto A, Greene MC, Powell BJ, et al. Application of the Expert Recommendations for Implementing Change (ERIC) compilation of strategies to health intervention implementation in low- and middle-income countries: a systematic review. Implementation science : IS. 2023 Oct 30;18(1):56. PubMed PMID: 37904218. PMCID: PMC10617067. Epub 2023/10/31.

33. Wendland, CL. Partial stories: Maternal death from six angles. University of Chicago Press; 2022 Apr 22.

34. Whyte SR, Whyte MA, Meinert L, Twebaze J. Therapeutic Clientship Belonging in Uganda’s Projectified Landscape of AIDS Care. In: Biehl JG, Petryna A, editors. When people come first: critical studies in global health. Princeton. Course Book ed. Princeton: Princeton University Press; 2013.

35. Londoño E, Gupta R, Stuyft PVd, Heine M, Giraldo G, Ku GM, et al. HEARTS quality: a policy framework to strengthen hypertension and cardiovascular risk management in primary healthcare—insights from HEARTS in the Americas. The Lancet Regional Health - Americas. 2026;53.

